# Global research trend in the treatment of the new Coronavirus diseases (COVID-19): bibliometric analysis

**DOI:** 10.1101/2020.06.13.20122762

**Authors:** Maxime Descartes Mbogning Fonkou, Abdourahamane Yacouba

## Abstract

The Coronavirus 2019 (COVID-19) pandemic has caused worldwide concern and has become a major medical problem. Vaccines and therapeutics are important interventions for the management of this outbreak. This study aims to used bibliometric methods to identify research trends in the domain of therapeutics and vaccines to cure patients with COVID-19 since the beginning of the pandemic.

The Web of Science Core Collection database was retrieved for articles on therapeutic approaches to coronavirus disease management published between January 1, 2020 and May 20, 2020. Identified and analyzed the data included title, corresponding author, language, publication time, publication type, research focus.

A total of 1569 articles on coronavirus therapeutic means from 84 countries were published in 620 journals. We note the remarkable progressive increase in the number of publications related to research on therapies and vaccines for COVID-19. The United States provided the largest number of articles (405), followed by China (364). Journal of Medical Virology published most of them (n = 40). 1005 (64.05%) were articles, 286 (18.23%) were letters, 230 (14.66%) were reviews. The terms “COVID- 19” or “SARS-CoV-2” or “Coronavirus” or “hydroxychloroquine” or “chloroquine” or “2019-nCOV” or “ACE2” or “treatment” or “remdesivir” or “pneumonia” were most frequently used, as shown in the density visualization map. A network analysis based on keyword co-occurrence revealed five distinct types of studies: clinical, biological, epidemiological, pandemic management, and therapeutics (vaccines and treatments).

COVID-19 is a major disease that has had an impact on international public health at the global level. Several avenues for treatment and vaccines have been explored. Most of them focus on older drugs used to treat other diseases that have been effective for other types of coronaviruses. There is a discrepancy in the results obtained from the studies of the drugs included in this study. Randomized clinical trials are needed to evaluate older drugs and develop new treatment options.

## Introduction

The world has been facing the emergence of a new respiratory disease in Wuhan, China, since December 31, 2019. A few days later, Chinese scientists identified the causative virus, as novel coronavirus[1], and was subsequently named SARS-CoV-2 by the *Coronaviridae* Study Group (CSG) of the International Committee on Taxonomy of Viruses[2]. Since, global efforts focused on fighting Coronavirus disease 2019 (COVID-2019) pandemic. The high infectiousness of the virus posed a problem in countries where barrier measures were not being met and health care facilities became overcrowded and unable to accommodate the excess of patients. Up to now, the world health organization (WHO) and United States Centers for Disease Control and Prevention (CDC) guidelines for the management of COVID-19 have been limited to infection control and symptomatic management of patients[3,4]. There are no specific antiviral drugs or vaccines available for coronavirus. Doctors use different antiviral, antibiotic and anti-inflammatory agents based on expert opinion, case series and prospective and randomized trials conducted worldwide to relieve infected patients. It is against this background that we have referred to highlight the various treatments that have had a considerable impact on the reduction and, in particular, the spread of this pandemic throughout the world since its emergence.

## Methods

### Sources of Data and Search Strategy

All data were retrieved from all available journals of the Thomson Reuters Web of Knowledge on May 20, 2020 using the following search terms: (Treatment OR Vaccine) AND (COVID-19). The Web of Science provides comprehensive publication data and is the widely accepted and frequently used database for the analysis of scientific publications. The publication period considered in this study was from January 1, 2020 to May 20, 2020; a total of 4,330 articles were identified. All articles with focus on the treatment of the COVID-19 and published in 2020 were included. All age groups were included. Articles related to *Severe acute respiratory syndrome coronavirus* (SARS-CoV or SARS-CoV-1) or *Middle East respiratory syndrome-coronavirus* (MERS-CoV) were excluded. Moreover, articles focusing on the infection or physiopathology of the COVID-19 were excluded. Thus, a total of 1,569 articles were ultimately included in the final analyses after duplicated removed.

### Data Analysis

Publications were stratified and systematically assessed according to publication days, country, journal, and authors. Additionally, the frequencies of keywords extracted from the articles were assessed and then included in a network analysis. All data were downloaded from the Web of Science and imported into VOSviewer v.1.6.15 (Centre for Science and Technology Studies, Leiden University, Leiden, The Netherlands), which is commonly used to analyze and visualize relationships among authors, countries, co-citations, and the terms used in articles [5,6]. The visualization of similarities (VOS) mapping method was used to estimate similarity (affinity) according to association strength, where higher association strength is indicative of greater similarity between terms, and a larger number of publications in which two items co-occur indicates that the terms are more closely similar to each other. The number of clusters can be varied depending on threshold of similarity between the nodes. The resolution of clustering was set as the default value (1.00) in this study[5].

Additionally, keywords were analyzed to identify popular topics in research on treatments and vaccine to control COVID-19. Keywords indicate article research themes; co-occurring keywords reveal associations in underlying themes among articles. The VOS method was applied to cluster keywords into different groups; each cluster was identified with a different color. Each keyword is represented by a circle, the diameter and label size of which denote the number of occurrences. Colors represent groups of linked terms; the label size of a term represents the number of publications on acupuncture for pain control in which it is used, and the distance between two terms represents the degree to which they are associated.

## Results

### General information

The initial search yielded 4,330 preliminary results. After verifying the titles and abstracts of the articles obtained from these results and removed duplicated, we retained 1,569 articles that met our inclusion criteria (Figure 1). Types of retrieved documents are listed in Table 1. Original research articles (1,005; 64.05%) were the main type. A total of 12 different languages were encountered in the retrieved documents. English language (1,453; 92.61%) and Chinese (83; 5.29%) was most encountered languages Table 2. Figure 2 presents the distribution of publications on vaccine and treatment related to COVID-19 during the period of January 1–May 20, 2020. Since the first publication on 30 January, an average of 14 publications has been recorded each day.

**Figure 1:**
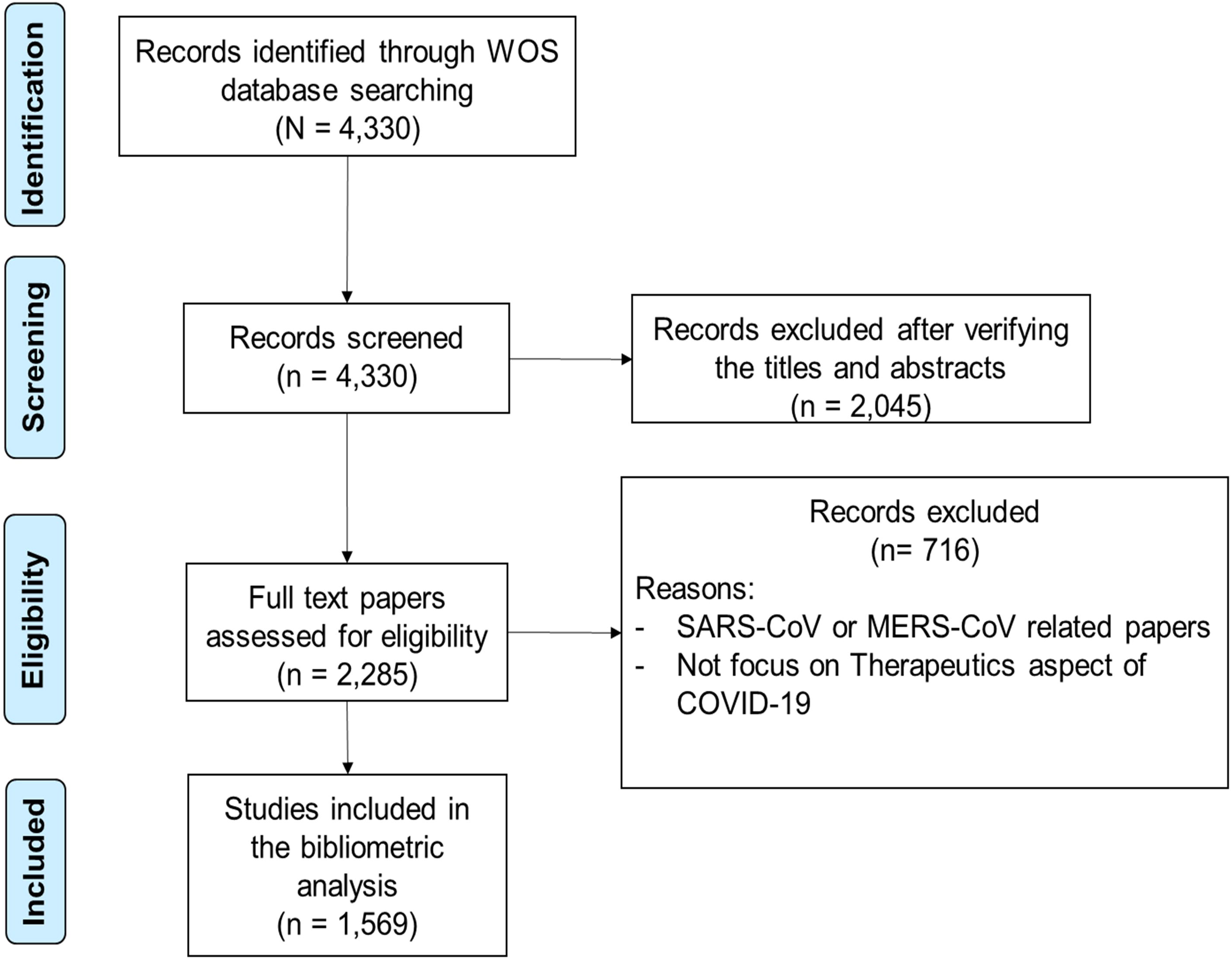
Flow chart of literature filtering included in this study.

**Table 1.** Types of retrieved documents

| Document Types | Records | % of 1569 |
| --- | --- | --- |
| Article | 1005 | 64.05 |
| Letter | 286 | 18.23 |
| Review | 230 | 14.66 |
| Editorial | 153 | 9.75 |
| Other | 122 | 7.78 |
| Early Access | 73 | 4.65 |
| News | 47 | 3.00 |
| Case Report | 34 | 2.17 |
| Abstract | 26 | 1.66 |
| Clinical Trial | 4 | 0.26 |
| Correction | 4 | 0.26 |
| Reference Material | 3 | 0.19 |
| Unspecified | 3 | 0.19 |
| Biography | 2 | 0.13 |

**Table 2.** Languages

| Languages | Records | % Of 1569 |
| --- | --- | --- |
| English | 1453 | 92.61 |
| Chinese | 83 | 5.29 |
| German | 10 | 0.64 |
| Spanish | 10 | 0.64 |
| French | 5 | 0.32 |
| Hungarian | 4 | 0.26 |
| Italian | 3 | 0.19 |
| Dutch | 2 | 0.12 |
| Swedish | 2 | 0.12 |
| Danish | 1 | 0.06 |
| Icelandic | 1 | 0.06 |
| Portuguese | 1 | 0.06 |

**Figure 2:**
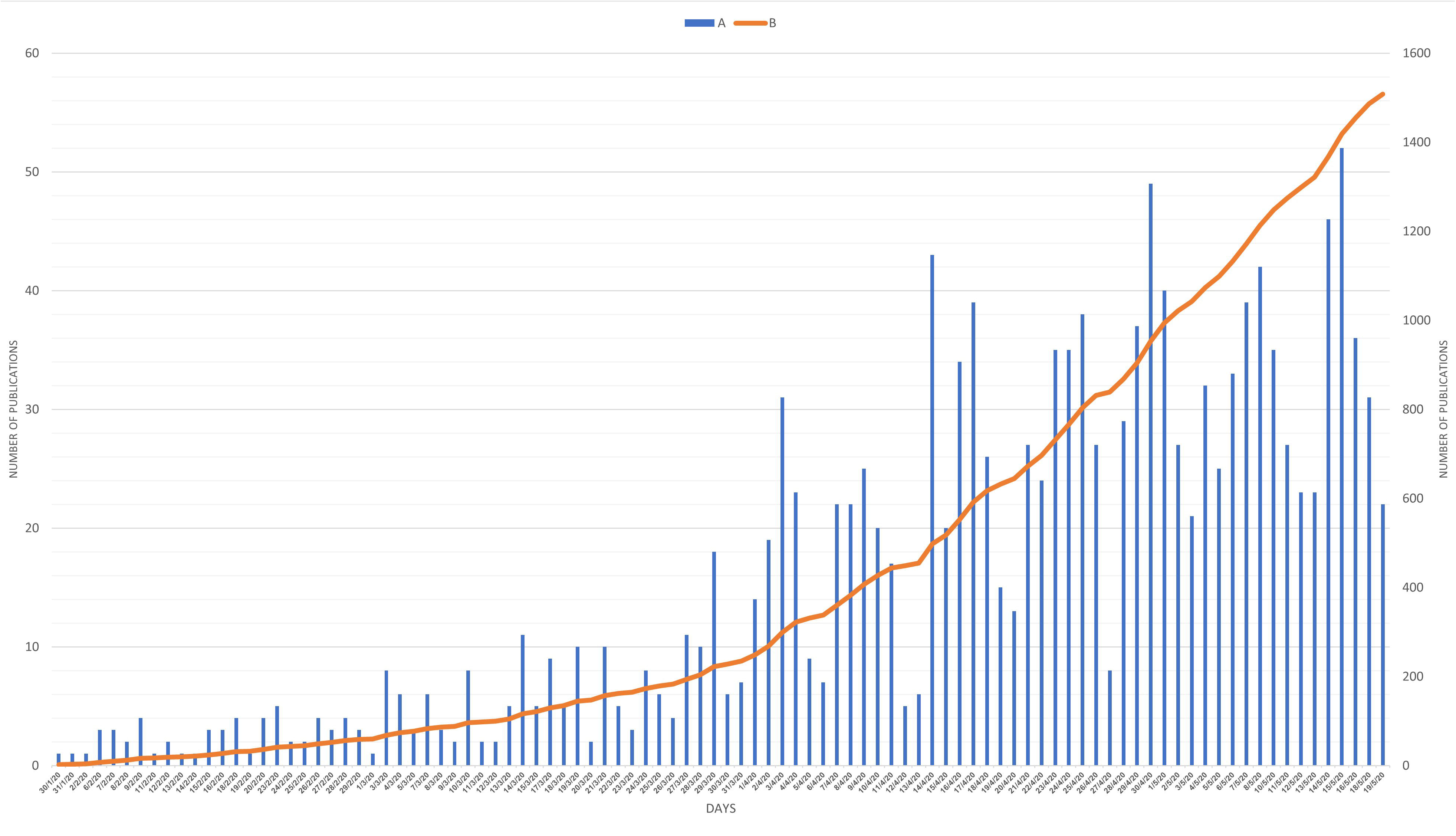
Number of publications per day that explore approaches to address the emerging coronavirus pandemic between January 1 and May 20, 2020. A-Number of publications per day; B- Cumulative publication

Of the 620 journals that were identified in the present study, *Journal of Medical Virology* published the most articles (40; 2.55%), followed by the *The New England Journal of Medicine* (27; 1.72%) and *International Journal of Antimicrobial Agents* (24; 1.53%) and *Nature* (24; 1.53%) (Table 3).

**Table 3.** Top 10 most productive journal

| Journal | Records | % Of 1569 | Journal Impact 2018 | H Index |
| --- | --- | --- | --- | --- |
| *Journal of Medical Virology* | 40 | 2.549 % | 2.049 | 105 |
| *The New England Journal Of Medicine* | 27 | 1.721 % | 37.909 | 933 |
| *Nature* | 24 | 1.530 % | 43.070 | 1096 |
| *International Journal of Antimicrobial Agents* | 24 | 1.530 % | 4.615 | 111 |
| *Journal of Biomolecular Structure Dynamics* | 23 | 1.466 % | 3.310 | 58 |
| *Annals of The Rheumatic Diseases* | 22 | 1.402 % | 14.299 | 212 |
| *Pharmacological Research* | 22 | 1.402 % | 5.574 | 118 |
| *Medical Hypotheses* | 21 | 1.338 % | 1.322 | 77 |
| *BMJ Clinical Research Ed* | 18 | 1.147 % | 0.48 | 0 |
| *Dermatologic Therapy* | 15 | 0.956 % | 1.74 | 60 |
| *European Review For Medical And Pharmacological Sciences* | 15 | 0.956 % | 2.721 | 48 |
| *Chinese Journal Of Tuberculosis And Respiratory Diseases* | 15 | 0.956 % | 0.340 | 16 |

### Geographical distribution of retrieved publications

Eighty-four countries have contributed to the search for therapeutics and vaccines to cure patients with COVID-19. Geographical distribution of retrieved publications was presented in world map using Microsoft Excel for Microsoft 365 MSO (version 16.0.12730.20144) (Figure 3). Top countries that participated in publishing documents on the therapeutics and vaccines to cure patients with COVID-19 were listed in Table 4. The United States of America (USA) was the most productive country in this field with 407 publications, follow by China 364 publications. The USA and China participated in half (49.47%) of worldwide productivity. Moreover, these gains are concentrated in countries with higher GDP and higher rate of patient’s death from COVID-19. Analysis of country coauthorships using VOSviewer showed a map with four clusters (Figure 4). Countries in the same cluster have higher collaboration than those distantly located in other clusters. Furthermore, countries with higher number of coauthorships had higher number of articles published on international collaboration. International cooperation analysis shows that the most frequent cooperation occurs in the United States and China (Figure 4).

**Figure 3:**
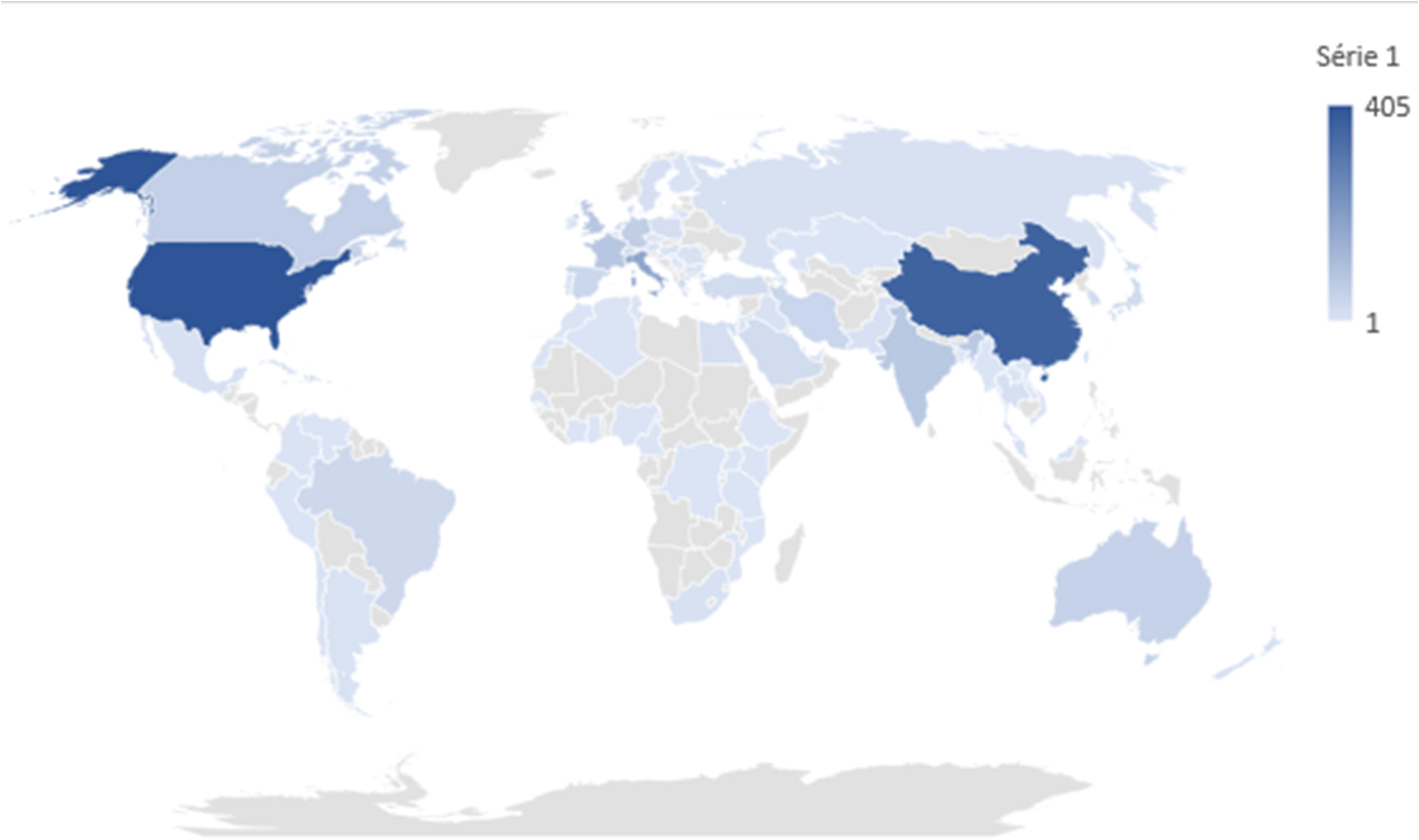
World maps showing the geographical distribution of retrieved publications.

**Table 4.** Top 15 most productive countries

| iso_code | Location | Total documents | Total confirmed COVID-19 cases | Total confirmed COVID-19 deaths | Total cases per million | Total deaths per million | GDP per capita | Population |
| --- | --- | --- | --- | --- | --- | --- | --- | --- |
| USA | United States | 405 | 1528568 | 91921 | 4617,993 | 277,705 | 54225,446 | 331002647 |
| CHN | China | 364 | 84065 | 4638 | 58,406 | 3,222 | 15308,712 | 1439323774 |
| ITA | Italy | 177 | 226699 | 32169 | 3749,457 | 532,055 | 35220,084 | 60461828 |
| GBR | United Kingdom | 84 | 248818 | 35341 | 3665,233 | 520,593 | 39753,244 | 67886004 |
| FRA | France | 83 | 143427 | 28022 | 2197,323 | 429,301 | 38605,671 | 65273512 |
| IND | India | 78 | 106750 | 3303 | 77,355 | 2,393 | 6426,674 | 1380004385 |
| DEU | Germany | 64 | 176007 | 8090 | 2100,725 | 96,558 | 45229,245 | 83783945 |
| CAN | Canada | 58 | 79101 | 5912 | 2095,826 | 156,642 | 44017,591 | 37742157 |
| AUS | Australia | 51 | 7068 | 99 | 277,178 | 3,882 | 44648,71 | 25499881 |
| IRN | Iran | 42 | 124603 | 7119 | 1483,494 | 84,757 | 19082,62 | 83992953 |
| ESP | Spain | 39 | 232555 | 27888 | 4973,93 | 596,474 | 34272,36 | 46754783 |
| CHE | Switzerland | 37 | 30535 | 1613 | 3528,174 | 186,374 | 57410,166 | 8654618 |
| BRA | Brazil | 34 | 271628 | 17971 | 1277,892 | 84,546 | 14103,452 | 212559409 |
| JPN | Japan | 27 | 16385 | 771 | 129,55 | 6,096 | 39002,223 | 126476458 |
| ISR | Israel | 24 | 16650 | 277 | 1923,623 | 32,003 | 33132,32 | 8655541 |
| OWID_WRL | World | 1569 | 4861974 | 323156 | 623,746 | 41,458 | 15469,207 | 7794798729 |

**Figure 4:**
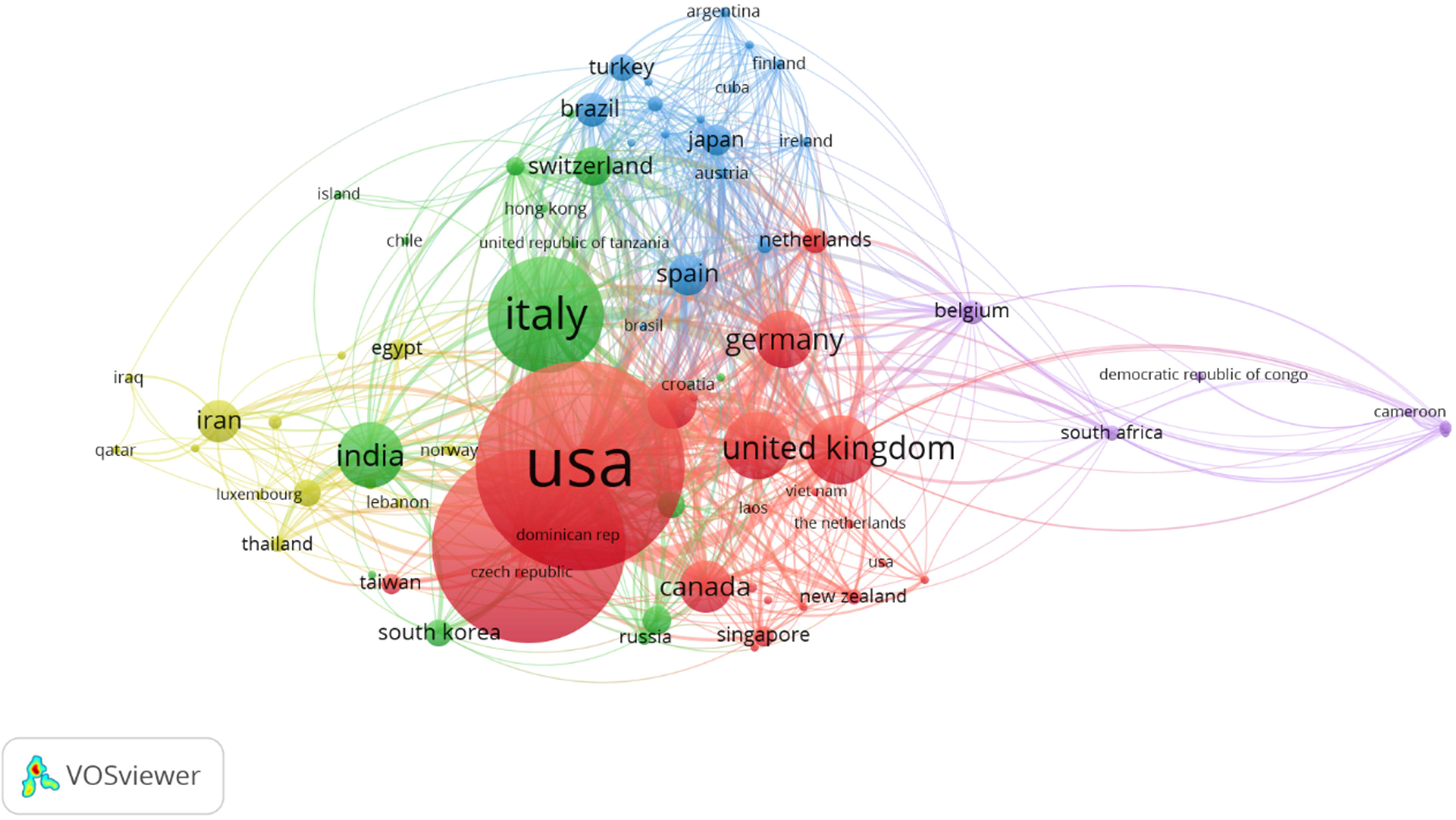
Analysis of country co-authorships. The node’s diameter is proportional to the country production. It appears that scientific publications were mainly driven by the research hubs such as the United States, China, Italy, United Kingdom, France, and India, which were also heavily hit by COVID-19.

### Profiling of authors

Of all 7,374 authors included in this subject, 774 authors published more than 2 papers and 17 authors published more than 5 papers. Table 5 shows the top 15 authors, according to the author lists of the articles included in the present study. Author’s analysis showed that 58 authors appeared more than 4 times (out of 7374 authors); these authors were classified into 21 clusters with 7 clusters with at least 3 authors. The most productive authors were clustered together in cluster numbers 1 and 2 mainly (Figure 5).

**Table 5.** Top 12 most productive authors

| Author |  | Documents | h-index | Average citations per item | Sum of Times Cited | Without self citations |
| --- | --- | --- | --- | --- | --- | --- |
| Jiang, Shibo | [1] Fudan Univ, Sch Basic Med Sci, Key Lab Med Mol Virol MOE NHC CAMS, Shanghai 200032, Peoples R China; [ 2 ] New York Blood Ctr, Lindsley F Kimball Res Inst, New York, NY 10065 USA | 11 | 5 | 8,27 | 91 | 83 |
| Rolain, Jean-Marc | Aix Marseille Univ, MEPHI, AP HM, IRD, IHU Mediterranee Infection, Marseille, France | 11 | 4 | 14,36 | 158 | 152 |
| Raoult, Didier | Aix Marseille Univ, MEPHI, AP HM, IRD, IHU Mediterranee Infection, Marseille, France | 10 | 4 | 15,6 | 156 | 151 |
| Colson, Philippe | Aix Marseille Univ, MEPHI, AP HM, IRD, IHU Mediterranee Infection, Marseille, France | 8 | 4 | 19,5 | 156 | 152 |
| Du, Lanying | Lindsley F. Kimball Research Institute, New York Blood Center, New York, NY, USA. | 8 | 4 | 6,63 | 53 | 48 |
| Mahase, Elisabeth | BMJ Publishing Group, British Med Assoc House, Tavistock Square, London Wc1h 9jr, England | 7 | 1 | 0,43 | 3 | 2 |
| Montecucco, Carlomaurizio | Rheumatology, Fondazione IRCCS Policlinico San Matteo, Pavia, Lombardia, Italy. | 7 | 1 | 0,33 | 2 | 2 |
| Shi, Zhengli | Chinese Acad Sci, Wuhan Inst Virol, CAS Key Lab Special Pathogens & Biosafety, Wuhan, Peoples R China | 6 | 5 | 41,5 | 249 | 245 |
| Zhang, Wei | PLA 900th Hospital of Joint Service CorpsFuzhou, Chinaand. | 6 | 4 | 7,71 | 54 | 54 |
| Baden, Lindsey R | the *New England Journal of Medicine* (NEJM) | 6 | 2 | 1,83 | 11 | 11 |
| Rubin, Eric J | the *New England Journal of Medicine* (NEJM) | 6 | 2 | 1,83 | 11 | 11 |
| Monti, Sara | Rheumatology, Fondazione IRCCS Policlinico S. Matteo, University of Pavia, Pavia, Italy Experimental Medicine, University of Pavia, Pavia, Italy" | 6 | 1 | 0,4 | 2 | 2 |

**Figure 5:**
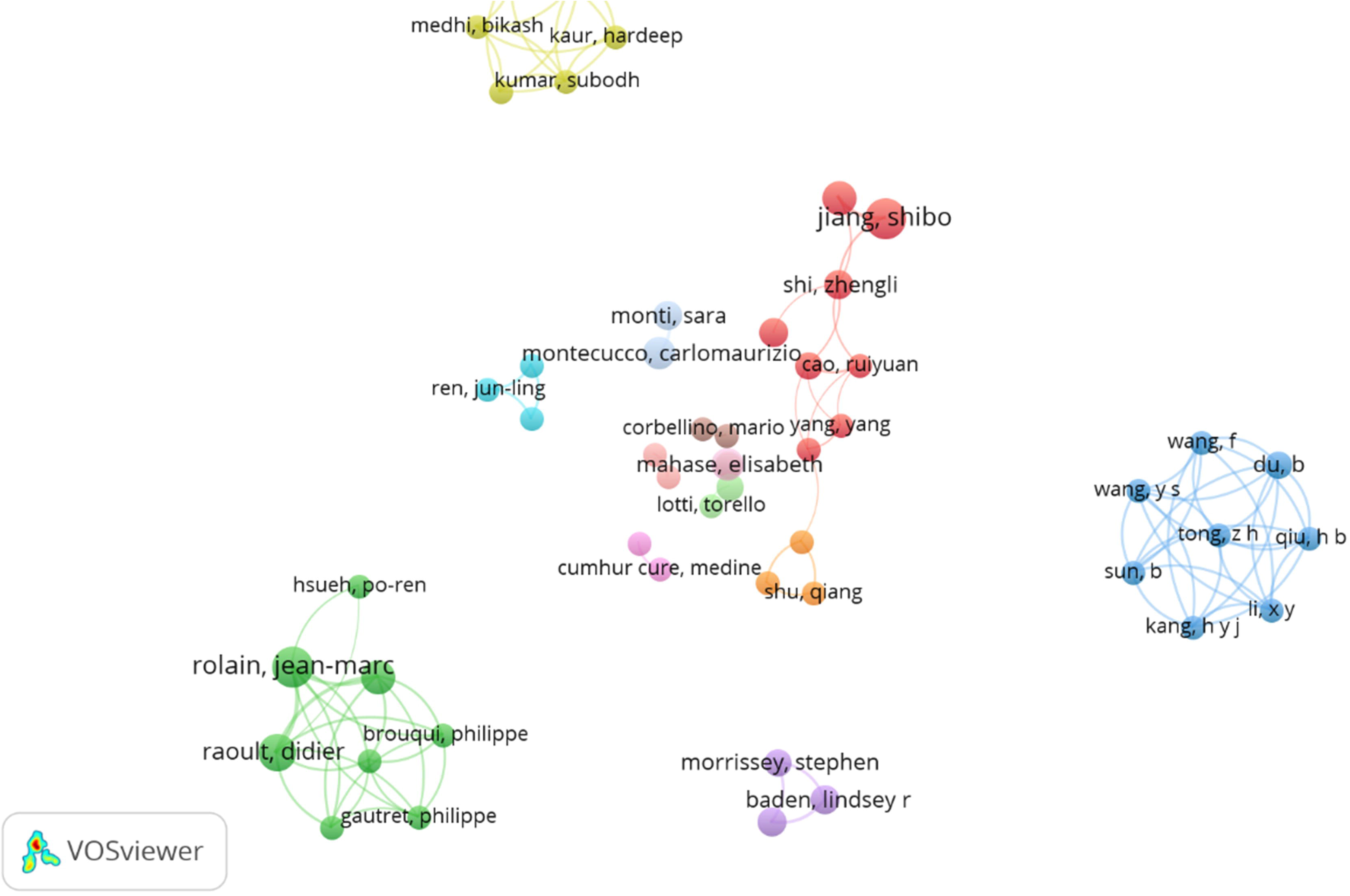
Network visualization map of study authors for researchers’ co-authorships. A minimum of 4 yielded 58 authors. The node’s diameter corresponds to the author production.

### Analysis of keywords

In this section, we investigated the keywords used in the therapeutic management of COVID-19 to discover the hotspot of this topic.

The counting of author keywords revealed that the most frequent treatment used as keywords were hydroxychloroquine (n = 78 repeats), followed by chloroquine (n = 56 repeats), remdesivir (n = 31 repeats), tocilizumab (n = 22 repeats), and azithromycin (n = 20 repeats).

To visualize the connection network between author keywords, we considered only keywords with at least 10 cooccurrences and found out that 38 of 1842 keywords were entered into the network and clustered into 4 groups (Figure 6).

**Figure 6:**
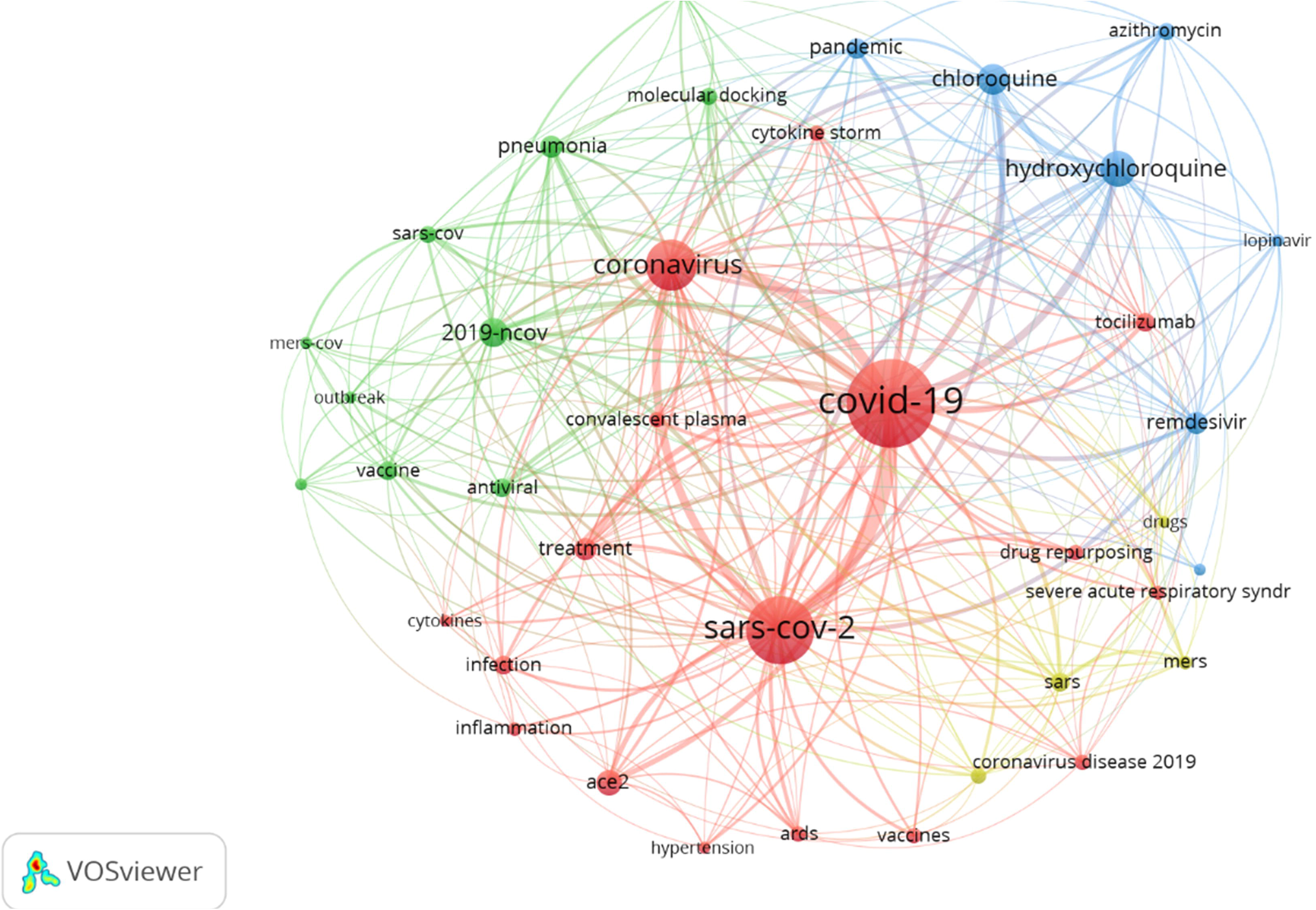
The connection network between author keywords with at least 10 co-occurrences. Out of 1842 keywords, 38 were entered into this network and clustered into 4 groups, which are depicted in different colors.

Cluster 1 (red) related to the “coronavirus diseases and its clinical issues”, including terms related to coronavirus, diseases complication and treatment.

Cluster 2 (green) could be called “disease epidemiology”, and includes terms like 2019-nCOV, MERS-CoV, SARS-CoV, novel coronavirus, pneumonia, outbreak, vaccine, and traditional Chinese medicine. This cluster contains 2 major topics: a) the main term “the disease” which in addition to the above-mentioned terms, but also b) “treatment and vaccine” which contains terms like vaccine, traditional Chinese medicine, and antiviral.

Cluster 3 (blue) related to “COVID-19 drug treatment”, with terms like coronavirus diseases 2019, pandemic, chloroquine, hydroxychloroquine, lopinavir, ritonavir, and remdesivir.

Cluster 4 (yellow) with terms including drugs, MERS, SARS and therapy.

Similarly, the counting of keywords Plus revealed that the most cooccurrence drugs in the therapeutic management of COVID-19 documents are hydroxychloroquine (n = 49 repeats) and chloroquine (n = 45 repeats).

To visualize the connection network between author keywords, we considered only keywords with at least 10 cooccurrences and found out that 73 of 871 keywords were entered into the network and clustered into 4 groups (Figure 7).

**Figure 7:**
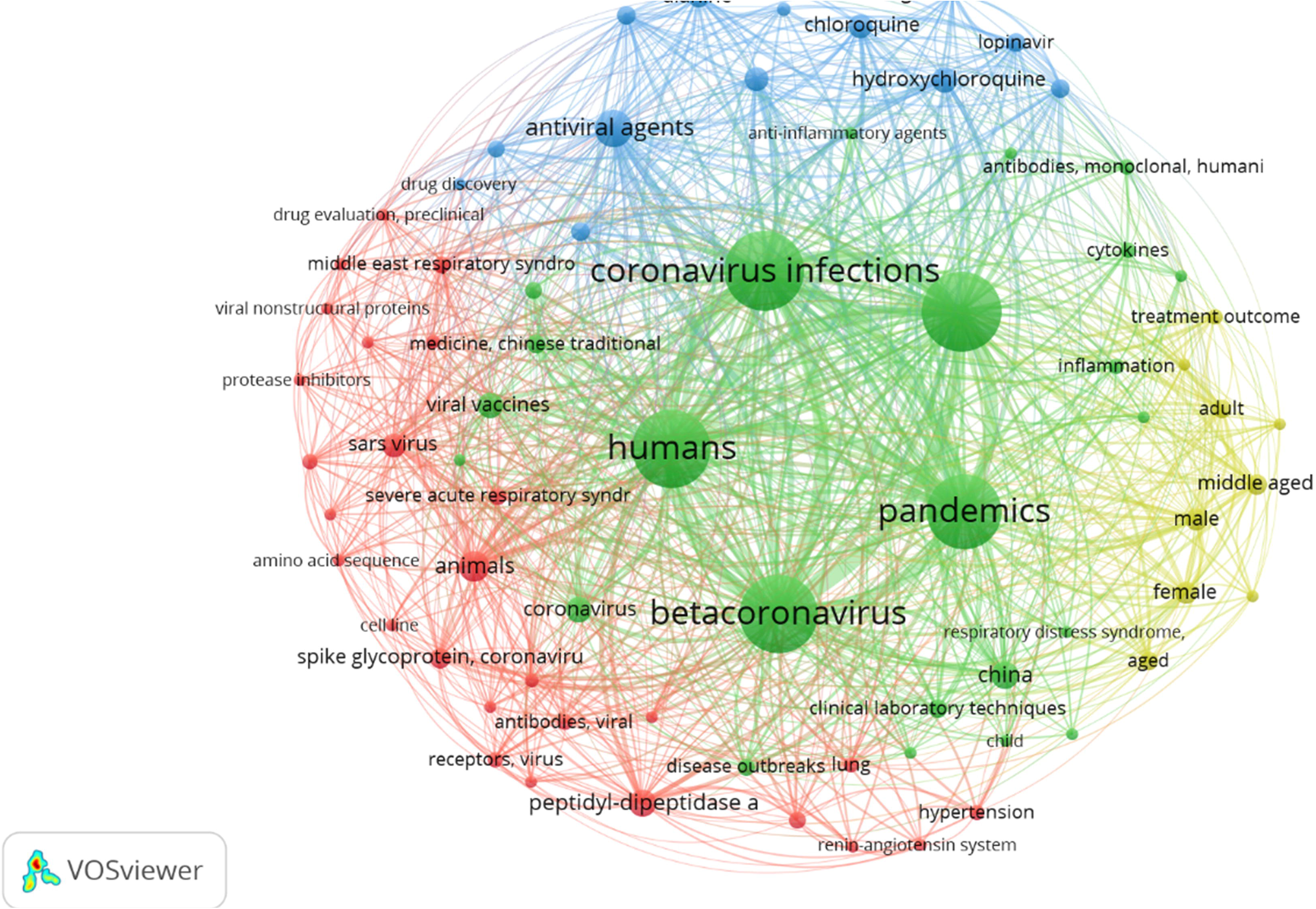
The connection network between keywords Plus with at least 10 co-occurrences. Out of 1842 keywords, 73 were entered into this network and clustered into 4 groups, which are depicted in different colors.

Cluster 1 (red) related to the “The virus and its biology”, including terms related to SARS virus and their biology. Within this cluster, we found the topic of “virus biology”, which included terms like glycoproteins, antibodies, viral structure, and viral replication.

Cluster 2 (green) relates to “The virus and pandemic management”, and include topics related to coronavirus and to pandemic, the disease outbreak and treatment management. Within this cluster, we found the topic of “care management”, which included terms on practice guidelines and clinical laboratory techniques. Further, within this cluster, we found the topic of “treatment management”, which included terms like viral vaccine, related to Chinese traditional medicine, and drug development.

Cluster 3 (blue) related to “drug treatment”, with terms like antiviral agents, chloroquine, hydroxychloroquine, lopinavir, ritonavir, drug repositioning, drug discovery and drug combinations.

Cluster 4 (yellow) related to “Clinical epidemiology”, with terms relating to age, gender, and procedures (like x-ray, CT, viral load).

### Most Influential Literature

Table 6 presents the 20 most cited articles in the field of therapeutic management of COVID-19. The number of citations ranged from 23 to 179, with an average citation per item of 55.6.

**Tableau 6.**
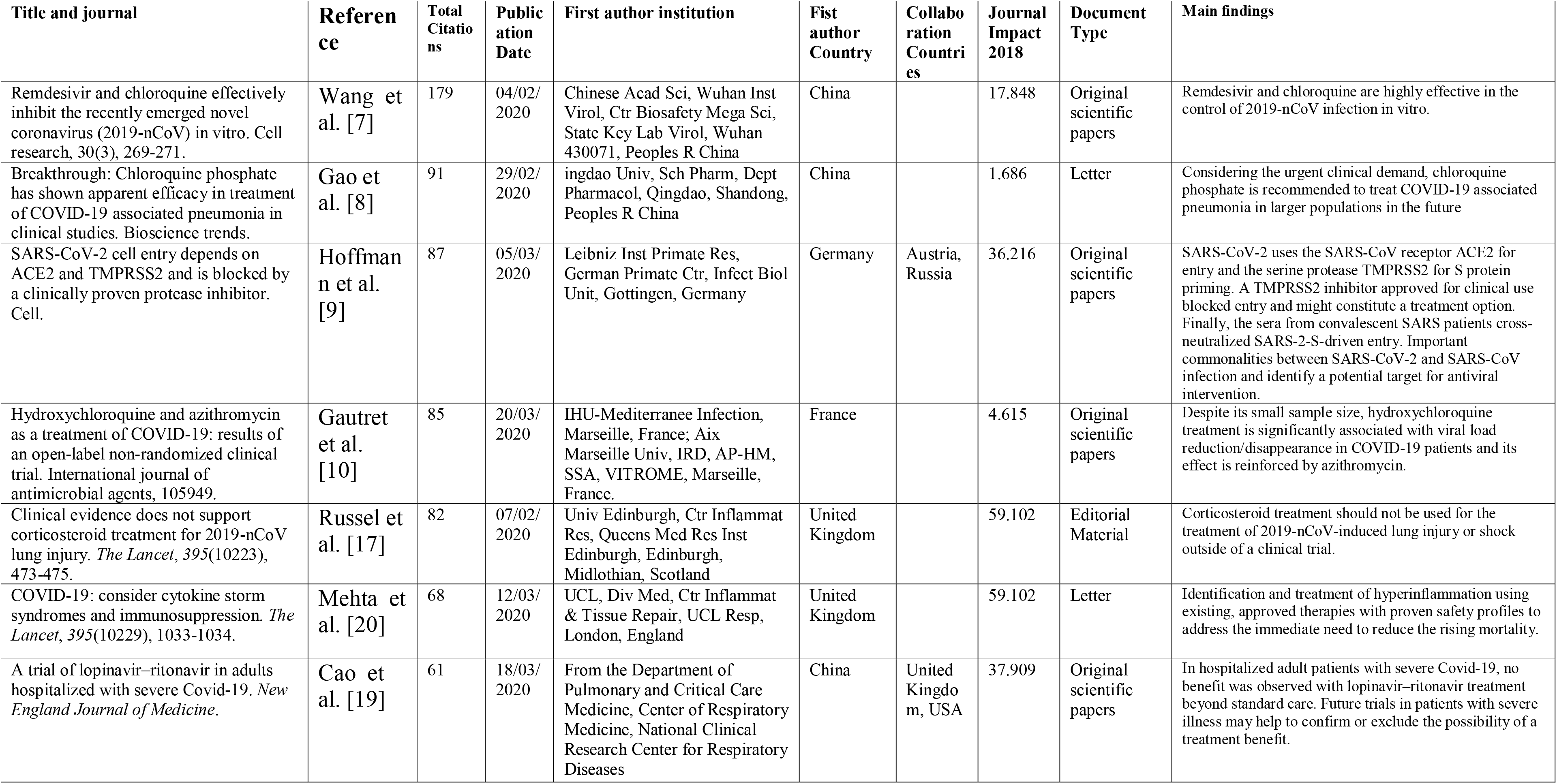
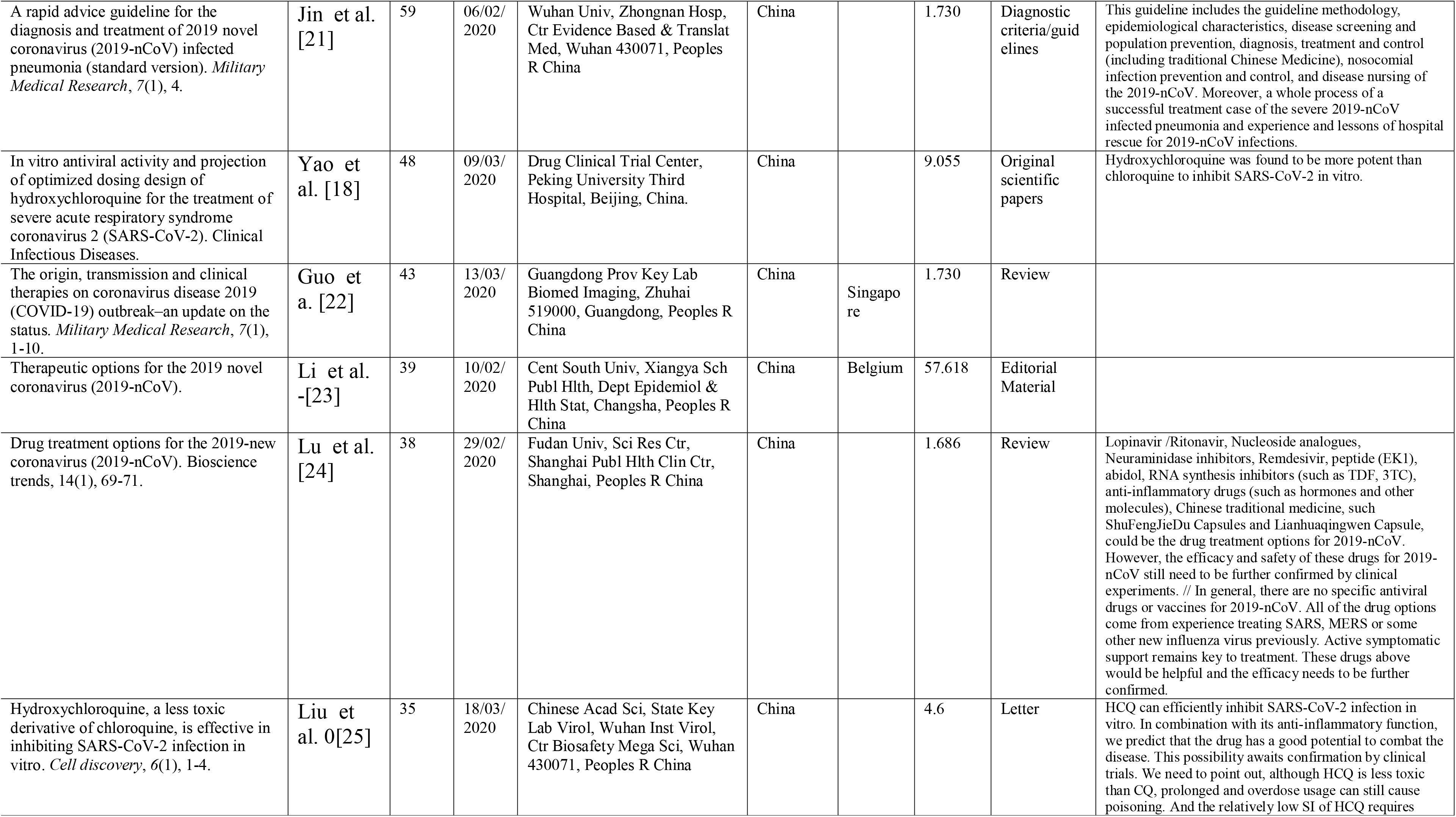
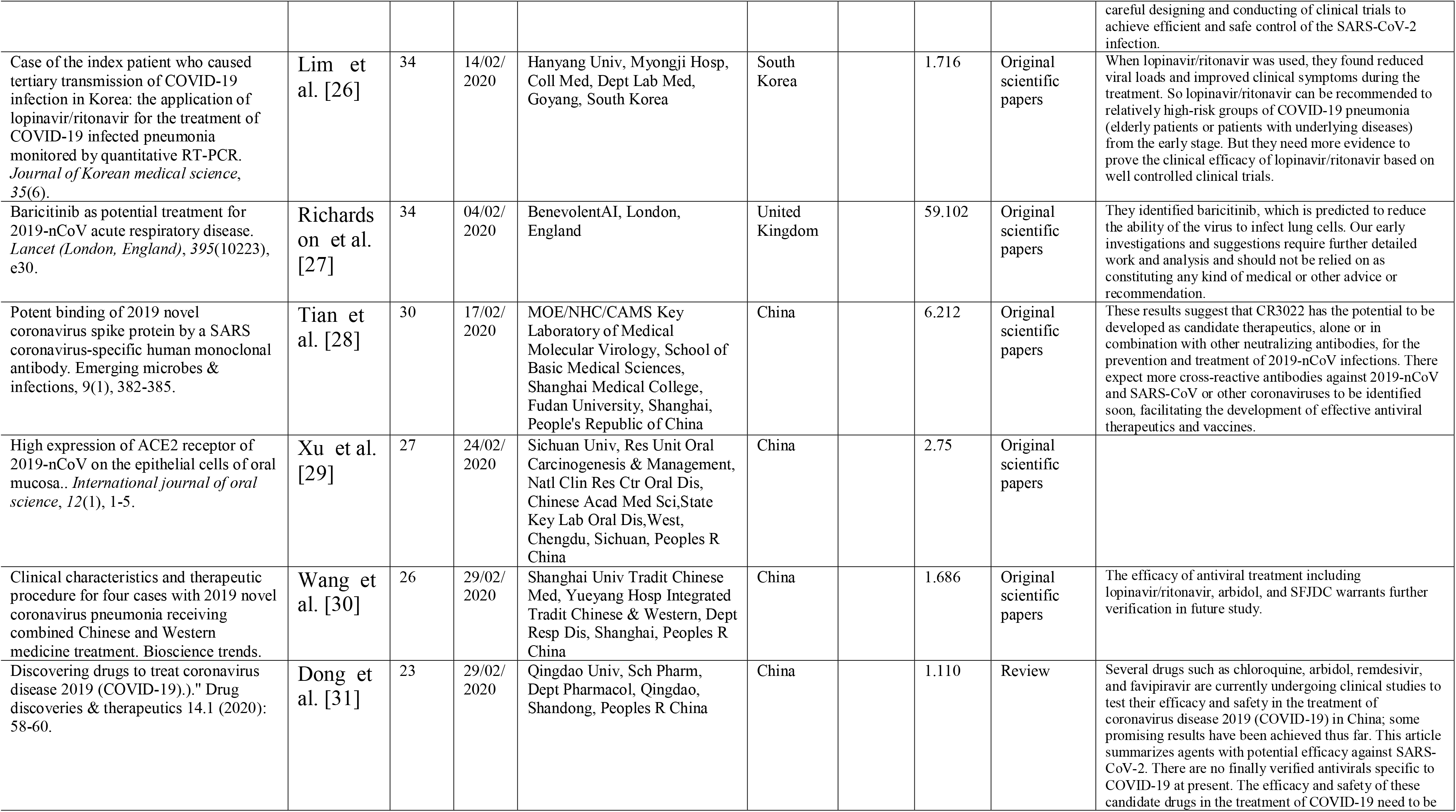
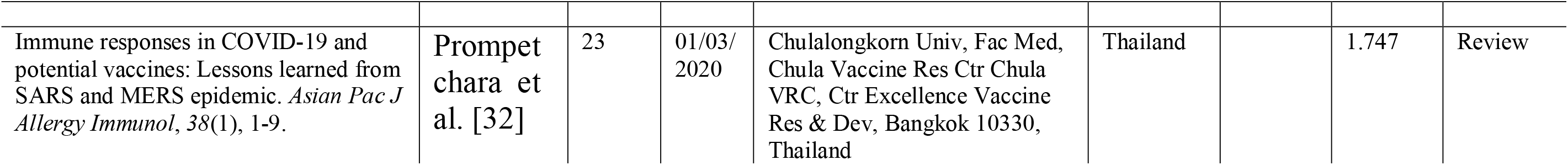
Top 20 most cited papers

These top 20 documents received the most citations among all documents (1112 times which comprise almost 47.76% of all citations). These documents consisted of 10 original articles, 3 letters, 4 reviews, 2 editorial, and 1 diagnostic criteria/guidelines. All the documents were published this year between February 4, to March 20, in 15 different journals including *The Lancet* (3 documents), *BioScience Trends* (3 documents) and *Military Medical Research* (2 documents).

The highest-ranking article was written by Wang *et al*. 2020 (*n* = 179 citations until May 23, 2020)[7], followed by a paper authored by Gao *et al*., 2020 (*n* = 91citations until May 23, 2020)[8], Hoffmann *et al*. 2020 (*n* = 87 citations until May 23, 2020)[9] and Gautret *et al*. 2020 (*n* = 85 citations until May 23, 2020) [10].

## Discussion

This bibliometric analysis of studies on treatment and vaccine to cure COVID-19 patients published since the beginning of the pandemic revealed that the total number of articles has steadily increased significantly since April. An average of 14 papers has been published per day since the first publication. An increase in research output has also been shown in similar research related to COVID-19 [11–13].

The largest proportion of articles in this area came from developed countries, including United States, followed by China, Italy, the United Kingdom and France. These countries, excluding China, are among the five countries with the highest number of deaths from COVID-19 on 20 May 2020 (https://covid19.who.int/). Possible explanations for these findings may be rapid economic growth or the progress of scientific research systems in these countries. These findings were like those reported in earlier bibliometric studies [11–15], which found that the economic growth of a country affected the quantity of research published by its researchers.

Keywords are chosen to reflect the main purpose of the entire study, in accordance with established research principles. They can provide information about the core content of an article and can also be used to identify research trends in a particular domain. Therefore, VOSviewer was used to analyze keywords from the work selected in this study that could be considered useful in organizing and prioritizing future research on treatments and vaccines to cure patients with COVID-19. A network analysis based on keyword co-occurrence revealed five distinct types of studies, namely clinical studies, biological studies, epidemiological studies, pandemic management, and studies related to vaccines and therapeutics.

The influential articles identified in this study explored several approaches to combat the new coronavirus pandemic. Guo, Yan-Rong et al In their review publish and cited more than 43 times, discussed the current treatment and scientific advances to combat the new coronavirus pandemic. Lopinavir/ritonavir, chloroquine, remdesivir (GS-5734), nafamostat, ribavirin, oseltamivir, penciclovir/ acyclovir, ganciclovir, favipiravir (T-705), and nitazoxanide have been listed as coronavirus treatment strategies, based on experience with the SARS-CoV and MERS-CoV epidemics. Rescue treatment with convalescent plasma and immunoglobulin G are delivered to some critical cases according to their conditions [16]. Russell et al 2020 [17] in a meta-analysis found that there is no clinical evidence that corticosteroids provide a net benefit in treating respiratory infections due to RSV, influenza, SARS-CoV or MERS-CoV and further conclude that corticosteroid treatment should not be used for the treatment of lung injury or shock induced by 2019-nCoV outside of a clinical trial[17]. The most widely cited article by Wang et al. 2020 [7] on 4 February, evaluated in vitro the antiviral efficacy of several FDA-approved drugs, including ribavirin, penciclovir, nitazoxanide, nafamostat, chloroquine, and two well-known broad-spectrum antiviral drugs, remdesivir (GS-5734) and favipiravir (T-705) against a clinical isolate of 2019-nCoV. The authors concluded that remdesivir and chloroquine are highly effective in controlling 2019-nCoV infection in vitro[7]. The second most cited paper, by Gao et al 2020[8] on February 19, recommends, given the urgent clinical demand, chloroquine phosphate to treat VID19-associated pneumonia in larger populations[8]. Another in vitro study conducted in China by Yao et al 2020 [18] on March 09, 2020 and cited nearly 50 times, shows that hydroxychloroquine was more potent than chloroquine in inhibiting SARS-CoV-2 in vitro[18]. Furthermore, despite the small sample size, Gautret et al 2020 [10]showed that hydroxychloroquine treatment is significantly associated with the reduction/disappearance of viral load in COVID-19 patients and that its effect is enhanced by azithromycin[10]. The study conducted by Cao et al 2020 [19] observed no benefit with lopinavir-ritonavir treatment beyond standard care by randomizing adult in patients with severe COVID-19 in a 1:1 ratio to receive either lopinavir-ritonavir in addition to standard care or standard care alone[19]. The third most cited article authored by Hoffmann et al 2020 [9] demonstrated that SARS-CoV-2 uses ACE2 receptor for entry and the serine protease TMPRSS2 for S protein priming. A TMPRSS2 inhibitor would block TMPRSS2 activity and might constitute a treatment option [9]

Our study has the advantage of being the first to provide a bibliometric overview of the search for therapeutic means to cure COVID-19. We have done our best to include all potential articles. Nevertheless, some limitations inherent to bibliometric analysis may be unavoidable. The database is continuously updated, and we have only selected literature from January 1, 2020 to May 20, 2020, without literature published after this date. The number of studies may increase rapidly with the breakthrough of future research. In addition, it appears that some medical journals are not indexed in Web of Sciences and therefore some articles may be missed. Some people may object to the use of Web of Sciences for data retrieval. However, we still believe that such a large amount of macroeconomic data and such a detailed analysis of the literature are sufficient to represent the trend of development in this area of research. Finally, we believe that researchers can ignore a minute number of potentially missing publications and that our results are sufficient to represent trends in this field.

## Conclusion

In conclusion, this study used bibliometric methods to identify research trends in the domain of therapeutics and vaccines to cure patients with COVID-19. The most important findings of this study are as follows: (1) the remarkable progressive increase in the number of publications related to research on therapies and vaccines for COVID-19 (2) most of these studies were conducted in the countries with the highest number of deaths (USA, Italy, United Kingdom and France) and China, first affected country (3) most of the publications were published in the Journal Of Medical Virology followed by The New England Journal Of Medicine and the International Journal Of Antimicrobial Agents (5) the terms “COVID- 19” or “SARS-CoV-2” or “Coronavirus” or “hydroxychloroquine” or “chloroquine” or “2019-nCOV” or “ACE2” or “treatment” or “remdesivir” or “pneumonia” were most frequently used, as shown in the density visualization map (6) A network analysis based on keyword cooccurrence revealed five distinct types of studies: clinical, biological, epidemiological, pandemic management, and therapeutics (vaccines and treatments).

## Data Availability

All data mentioned in the manuscript availability and will be immediately available on request.

## Acknowledgements

None

## Ethics approval and consent to participate

The analysis in this study is based on a retrospective bibliometric technique; therefore, no ethical approval was required.

